# Molecular point-of-care testing for influenza A/B and respiratory syncytial virus: comparison of workflow parameters for the ID Now and cobas Liat systems

**DOI:** 10.1101/19008227

**Authors:** Stephen Young, Jamie Phillips, Christen Griego-Fullbright, Aaron Wagner, Patricia Jim, Sheena Chaudhuri, Shaowu Tang, Joanna Sickler

**Affiliations:** University of New Mexico, Albuquerque, New Mexico, USA; TriCore Reference Laboratories, Albuquerque, New Mexico, USA; Roche Diagnostics Corporation, Indianapolis, Indiana, USA; Roche Molecular Systems, Inc., Pleasanton, California, USA

## Abstract

**Aims:** Point-of-care (POC) tests for influenza and respiratory syncytial virus (RSV) offer the potential to improve patient management and antimicrobial stewardship. Studies have focused on performance; however, no workflow assessments have been published comparing POC molecular tests. This study compared the Liat and ID Now systems workflow, to assist end-users in selecting an influenza and/or RSV POC test.

**Methods:** Staffing, walk-away, and turnaround time (TAT) of the Liat and ID Now systems were determined using 40 nasopharyngeal samples, positive for influenza or RSV. The ID Now system requires separate tests for influenza and RSV, so parallel (two instruments) and sequential (one instrument) workflows were evaluated.

**Results:** The ID Now ranged 4.1–6.2 minutes for staffing, 1.9–10.9 minutes for walk-away and 6.4–15.8 minutes for TAT per result. The Liat ranged 1.1–1.8 minutes for staffing, 20.0–20.5 minutes for walk-away and 21.3–22.0 minutes for TAT. Mean walk-away time comprised 38.0% (influenza positive) and 68.1% (influenza negative) of TAT for ID Now and 93.7% (influenza/RSV) for Liat. The ID Now parallel workflow resulted in medians of 5.9 minutes for staffing, 9.7 minutes for walk-away, and 15.6 minutes for TAT. Assuming prevalence of 20% influenza and 20% RSV, the ID Now sequential workflow resulted in medians of 9.4 minutes for staffing, 17.4 minutes for walk-away, and 27.1 minutes for TAT.

**Conclusions:** The ID Now and Liat systems offer different workflow characteristics. Key considerations for implementation include value of both influenza and RSV results, clinical setting, staffing capacity, and instrument(s) placement.

## INTRODUCTION

Respiratory tract infections are a major public health issue and significant cause of morbidity and mortality worldwide.[1] Influenza virus [2, 3] and respiratory syncytial virus (RSV) [4-6] are significant causes of respiratory illness and have overlapping clinical symptoms and seasonality.

Influenza is characterized by fever, cough, headache, muscle and joint pain, severe malaise, sore throat, and runny nose.[2] It can affect people of all ages, although some groups are at greater risk of severe disease or complications (young children, pregnant women, the elderly, individuals with chronic medical conditions, and those with immunosuppressive conditions).[2] Influenza virus types A and B are responsible for annual epidemics estimated to cause 3–5 million cases of severe illness and 290,000 to 650,000 deaths worldwide.[2] RSV is typically associated with mild symptoms in adults, including runny nose, cough, fever, and wheezing,[4] but is known to cause more serious illness in infants and older adults, including bronchiolitis and pneumonia.[4] RSV is a common cause of childhood acute lower respiratory infection, and was estimated, in 2015, to have caused 3.2 million hospital admissions and 59,600 in-hospital deaths in children younger than 5 years globally.[5] In the same year, there were an estimated 336,000 hospitalizations and 14,000 in-hospital deaths due to RSV-associated acute respiratory infection in older adults.[6]

However, in adults, RSV is likely to be under-reported as the symptoms are similar to those of influenza, making it “nearly impossible” to distinguish the two infections by clinical presentation alone.[7] A recent study assessing laboratory-confirmed hospitalized RSV cases in the US found that only 44% had RSV-specific International Classification of Diseases discharge codes (in the first nine positions), suggesting that administrative data underestimate adult RSV-related hospitalizations.[8] Furthermore, studies suggest that antibiotic use, morbidity, and mortality are higher among RSV-positive than influenza-positive adult hospitalized patients [9-11] and RSV patients are more likely to need hospital re-admission.[12]

Increased detection of RSV would be valuable, not only for appropriate management of patients but also to minimize transmission to high-risk groups. Reducing the time to detection of influenza and RSV could improve infection control and reduce in-hospital costs by improving the efficiency of patient triage and minimizing the time patients are kept in isolation while waiting for test results.[13] A fast and highly accurate diagnosis could also enable time-critical antivirals to be administered sooner [14] and reduce inappropriate antimicrobial administration.[15]

The recent introduction of point-of-care (POC) molecular tests for RSV and influenza A/B offers the potential for clinicians to have rapid nucleic acid amplification test (NAAT) results for these infections.[16, 17] These include the ID Now^™^ RSV assay and the ID Now^™^ Influenza A & B assay for use on the ID Now^™^ (formerly referred to as Alere^™^ i) system (ID Now), and the cobas^®^ Influenza A/B & RSV assay for use on the cobas^®^ Liat^®^ PCR System (Liat). To date, comparison studies have focused on performance based on the sensitivity and specificity of the ID Now and Liat assays.[18, 19] Differences between the two systems in terms of test operation or workflow have been mentioned but have not been the focus of the studies.[19, 20]

The two systems differ in terms of the manual steps involved, turnaround times (TAT), and multiplexing capabilities, which may impact choice of instrument for influenza and RSV testing. To further help clinicians and laboratory directors select their influenza and RSV testing methods, this study aimed to assess the workflow and ease of use of the ID Now and Liat systems. Specific features that were considered include TAT, staffing time, walk-away time, and the early call-out feature of the ID Now system.

## MATERIALS AND METHODS

### Design

Influenza A/B and RSV tests were run on the ID Now and Liat systems. For the ID Now system, sequential and parallel workflow modes were assessed because two tests (influenza A/B test and RSV test) are required to detect all three viruses. The tests were run by four trained operators, each working four 4-hour shifts (two shifts per day over two consecutive days). An independent time motion expert measured TAT and operator staffing and walk-away times. All operators spent at least 1 day using both systems before the ease of use and workflow testing to ensure the data captured routine use not initial use. All testing was conducted at TriCore Reference Laboratories (Albuquerque, New Mexico, USA) in November 2018.

### Samples

Forty positive residual de-identified nasopharyngeal samples, previously collected for routine clinical care, were used for this study. The samples were all positive for either influenza A, influenza B or RSV according to routine laboratory polymerase chain reaction (PCR) testing and had been frozen after all of the standard of care (SOC) testing was complete. Ten samples were provided for testing in a random order for each 4-hour shift (four positive for influenza A, two positive for influenza B, and four positive for RSV). Each sample was tested for all three viruses using the workflow modes described below. Sample volumes used were 200 μL for each ID Now assay (400 μL total) and 200 μL for the Liat influenza A/B and RSV assay. Two controls for each assay were run for each reagent lot.

### Assays and instruments

The testing used the ID Now Influenza A & B 2 assay and the ID Now RSV assay performed on the ID Now system (Abbott Diagnostics, Lake Forest, Illinois, USA) and the cobas Influenza A/B & RSV assay for use on the cobas Liat PCR System (Roche Molecular Diagnostics, Pleasanton, California, USA).

The ID Now system uses isothermal nucleic acid amplification technology and the Liat system uses PCR to detect specific nucleic acid sequences. The ID Now influenza A/B assay detects and differentiates influenza A and influenza B, and the ID Now RSV assay detects RSV. The Liat influenza A/B and RSV assay detects and differentiates influenza A, influenza B and RSV from a single sample. All three assays are Clinical Laboratory Improvement Amendments (CLIA)-waived *in vitro* diagnostic tests.[21-23]

### Workflow modes

The ID Now sequential workflow involved one operator running the ID Now influenza A/B assay followed by the ID Now RSV assay. The ID Now parallel workflow involved one operator running both ID Now assays at the same time on two separate instruments. Both workflows were evaluated to consider the different methods clinical sites may use to execute both tests. Each sample was run twice on the ID Now system, once in a parallel workflow and once in a sequential workflow shift. The Liat influenza A/B and RSV assay is a multiplex test that provides results for influenza A, influenza B and RSV from one sample run; therefore, multiple workflows were not required. However, each operator still ran through two Liat shifts so that they interacted equally with both systems.

### Analyses

TAT, staffing time and walk-away time (mean, median, and range) were determined for the ID Now and Liat systems for all workflows. TAT was the total time from the point when the initial set-up began to when the test result was released. Staffing time corresponded to the sum of the labor time (hands-on time) and oversight time (time that the operator was not directly engaged with the testing but had to remain close to the instrument). Walk-away time corresponded to the wait times when the operator was not required to be close to the instrument.

The ID Now system has an early call-out feature for positive test results. Therefore, for ID Now, TAT, staffing time, and walk-away time were calculated separately for positive and negative results. The early call-out feature does not guarantee that all positive results terminate early so waiting the full run time is still required. The Liat system does not have an early call-out feature and one test covers all three viruses, therefore, all results were combined for the Liat analyses.

The impact of early call-out on the average TAT, staffing and walk-away times in a real-world clinical setting was estimated by analyzing the expected median TAT, staffing and walk-away times based on different influenza and RSV prevalence levels.

Expected median time for prevalence ranging from 5% (0.05) to 30% (0.3) was calculated as follows:

> Prevalence expressed as a decimal*median time (TAT/staffing/walk-away) for a positive result + (1-prevalence expressed as a decimal)*median time (TAT/staffing/walk-away) for a negative result

### Ethics

The study was conducted in compliance with the International Conference on Harmonization Good Clinical Practice Guidelines, and applicable US Food and Drug Administration regulations. The nasopharyngeal specimens used in the study were de-identified by an honest broker using a Institutional Review Board-approved procedure.

## RESULTS

### Workflow steps per run

The ID Now system has eight manual steps (opening lid, inserting test base, inserting sample receiver, removing seal and inserting swab, transferring cartridge to receiver, transferring cartridge to test base, running test, discarding components) (Figure 1). The operator must wait for 3 minutes for the system to warm up before inserting the swab/sample between steps 3 and 4, calculated as staffing time in the analysis. The Liat system has four manual steps (scanning assay tube and inserting sample, close tube and scan, inserting assay tube and running test, discarding the tube).

**Figure 1.**
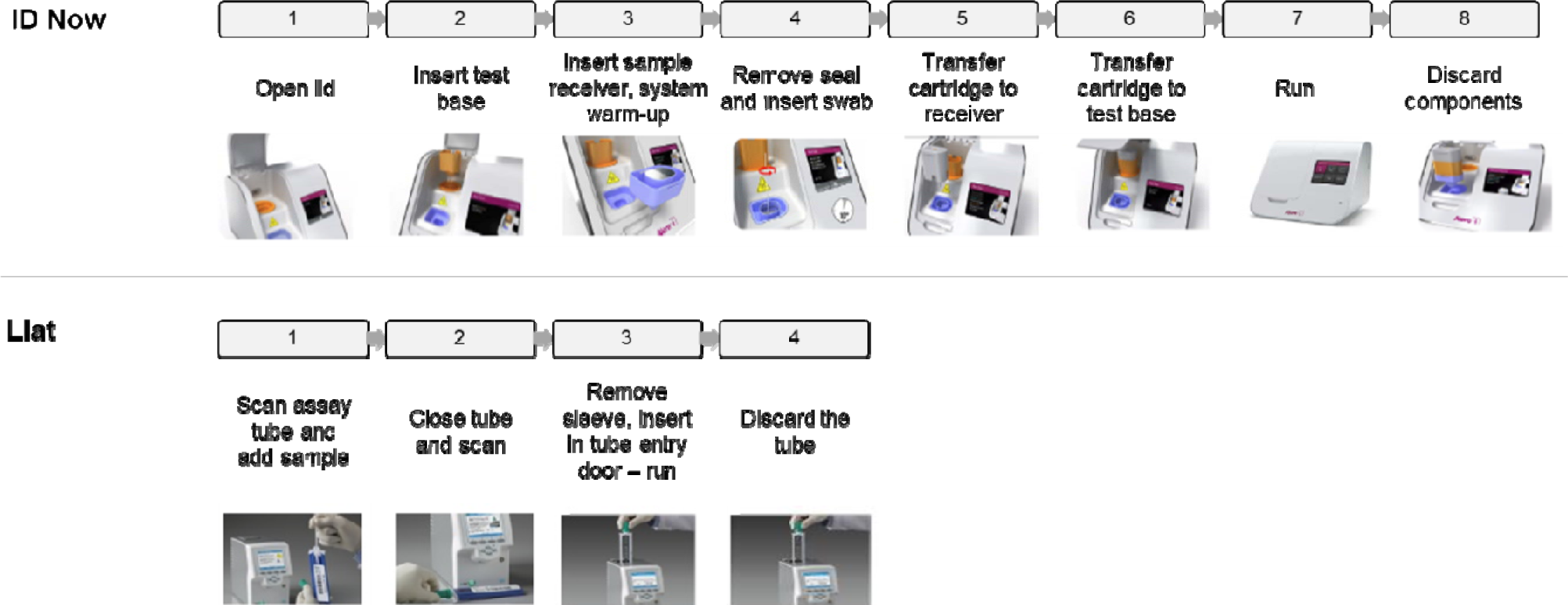
Workflow steps required to run a sample on the ID Now and Liat systems per run

### Workflow timings per run

The ID Now workflow timings varied per run by test type (influenza or RSV) and result, ranging 4.1–6.2 minutes for staffing time, 1.9–10.9 minutes for walk-away time and 6.4– 15.8 minutes for TAT per test (Table 1). The early call-out feature of the ID Now system was utilized for the study, which resulted in much shorter TAT and walk-away times for positive results than for negative results (Table 1 and Figure 2). For the ID Now assays, TAT ranged 6.4–9.6 minutes for positive results and 14.4–15.8 minutes for negative results, and walk-away time ranged 1.9–4.2 minutes for positive results and 10.0–10.9 minutes for negative results (Table 1). Overall staffing time for the ID Now assays was comparable for positive (4.1–6.2 minutes) and negative results (4.1–5.6 minutes) (Table 1), and 31.9–38% of the TAT for influenza runs (Figure 3a and 3b).

**Table 1.** Staffing time, walk-away time, and TAT per run

|  | **ID Now<sup>a</sup>** |  |  |  | **Liat<sup>b</sup>** |
| --- | --- | --- | --- | --- | --- |
|  | **Influenza A/B assay** |  | **RSV assay** |  | **Influenza A/B & RSV assay** |
|  | **Positive** | **Negative** | **Positive** | **Negative** | **Positive or negative** |
| **Number of tests** | 20 | 20 | 13 | 27 | 80 |
| **Staffing time (minutes)** |  |  |  |  |  |
| Mean | 4.82 | 4.82 | 4.68 | 4.72 | 1.36 |
| Median | 4.71 | 4.68 | 4.63 | 4.68 | 1.37 |
| Range | 4.17–6.20 | 4.35–5.60 | 4.13–5.60 | 4.08–5.23 | 1.07–1.77 |
| SD | 0.39 |  | 0.35 |  | 0.13 |
| **Walk-away time (minutes)** |  |  |  |  |  |
| Mean | 2.99 | 10.30 | 2.72 | 10.27 | 20.22 |
| Median | 2.88 | 10.23 | 2.67 | 10.18 | 20.20 |
| Range | 1.87–4.23 | 10.02–10.87 | 2.18–3.40 | 10.00–10.87 | 20.03–20.45 |
| SD | 3.73 |  | 3.59 |  | 0.10 |
| **TAT (minutes)** |  |  |  |  |  |
| Mean | 7.81 | 15.11 | 7.41 | 14.98 | 21.58 |
| Median | 7.69 | 15.11 | 7.47 | 14.92 | 21.57 |
| Range | 6.40–9.57 | 14.68–15.73 | 6.52–8.03 | 14.40–15.75 | 21.25–21.97 |
| SD | 3.74 |  | 3.61 |  | 0.15 |

<sup>a</sup>ID Now data are from sequential workflow runs
<sup>b</sup>Liat data are from all runs combined
RSV, respiratory syncytial virus; SD, standard deviation; TAT, turnaround time

**Figure 2.**
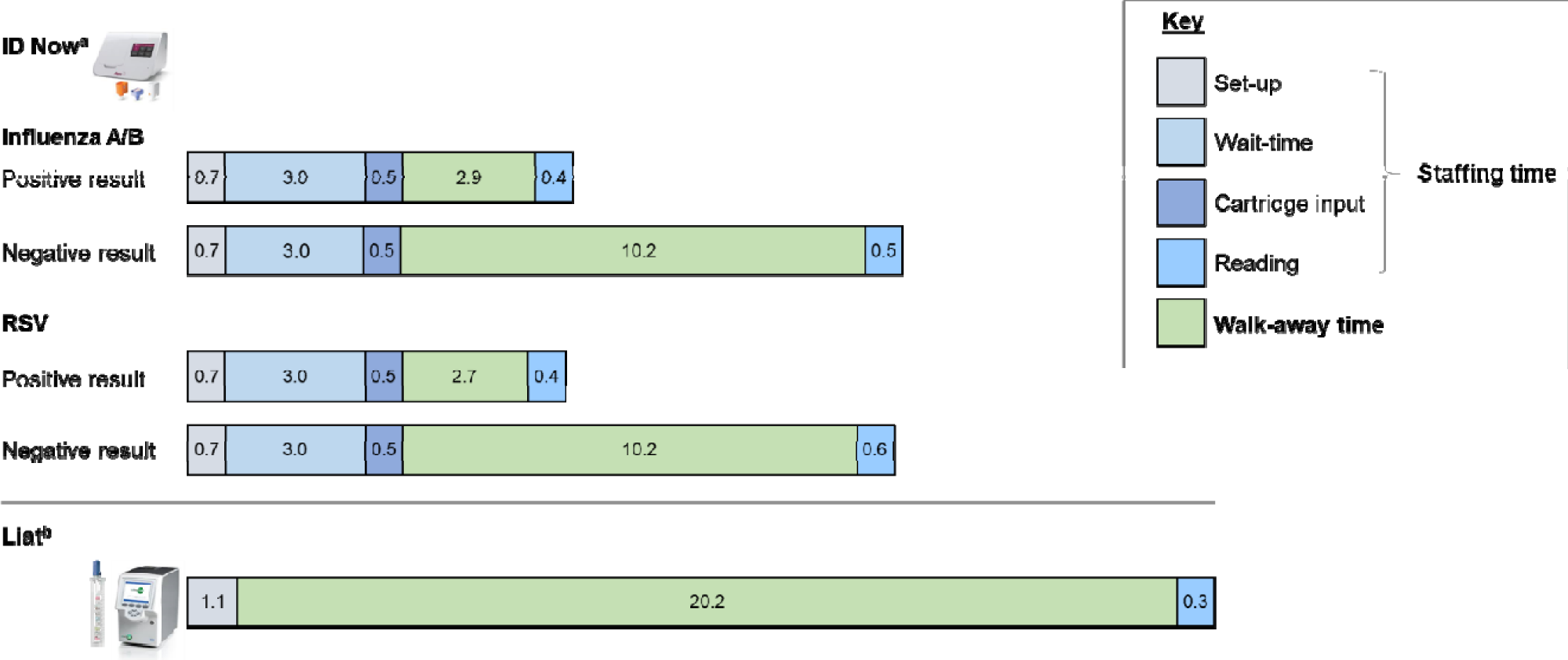
Breakdown of test timings (median times in minutes per run) ^a^ID Now data are from sequential workflow runs ^b^Liat data are from all runs combined RSV, respiratory syncytial virus

**Figure 3.**
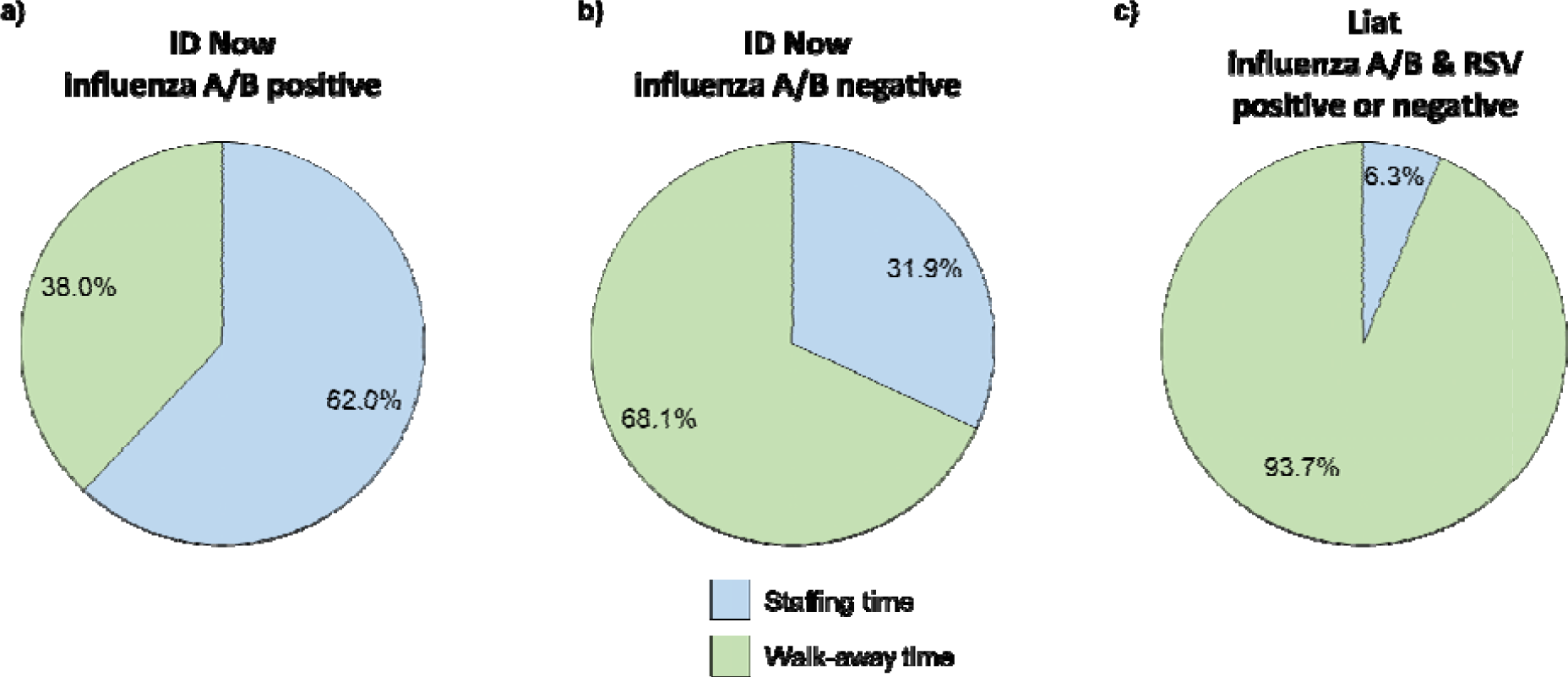
Mean staffing time and walk-away time as percentages of the TAT for a) ID Now positive influenza tests, b) ID Now negative influenza tests, and c) Liat positive or negative tests RSV, respiratory syncytial virus; TAT, turnaround time

The Liat ranged 1.1–1.8 minutes for staffing time, 20.0–20.5 minutes for walk-away time, and 21.3–22.0 minutes for TAT for influenza and RSV results (Table 1). The Liat combined influenza/RSV test and no early call-out feature made results consistent for all testing scenarios, with staffing time accounting for 6.3% of the TAT (Figure 3c).

### Workflow timings for influenza A/B and RSV

For the ID Now parallel workflow, the median (range) times for all influenza and RSV tests were 5.9 minutes (5.0–7.4 minutes) for staffing time, 9.7 minutes (9.4–11.1 minutes) for walk-away time, and 15.6 minutes (14.7–16.8 minutes) for TAT. However, for the ID Now system, the early call-out for positive results means that test results will influence TAT. When assessing ID Now negative results only, the parallel workflow median TAT was shorter (15.1 minutes) compared with the sequential workflow (30.0 minutes) (Figure 4).

**Figure 4.**
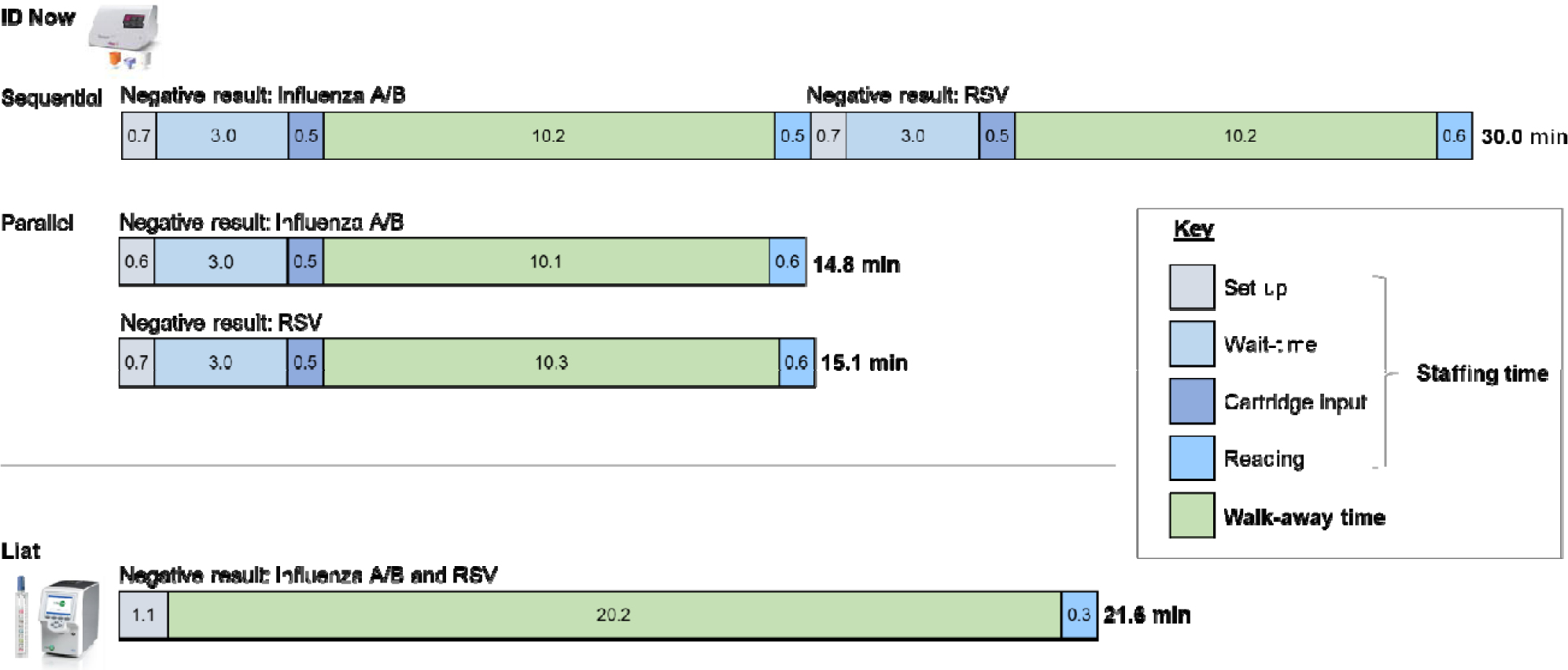
Breakdown of test timings for negative results (median times in minutes) for influenza A/B and RSV RSV, respiratory syncytial virus

For the ID Now sequential workflow in a real-world setting, the prevalence of influenza and RSV will be a key determinant of TAT. As a result, the data were used to make a theoretical calculation of TAT (Table 2), as well as staffing time and walk-away time (Table 3). Expected median TAT for the ID Now influenza A/B and RSV assays (sequential workflow) ranged from 25.6 minutes for a prevalence of 30% for each virus to 29.3 minutes for a prevalence of 5% for each virus (Table 2). The walk-away time for the same prevalence range was 16.0–19.7 minutes (Table 3). Staffing time remained consistent at 9.4 minutes for all prevalence ranges. For influenza alone, the median TAT was also calculated for the ID Now assay, which ranged from 12.9–14.7 minutes based on a prevalence range of 30–5% (Table 4). For the Liat influenza A/B and RSV assay, infection prevalence had no impact on expected median TAT, staffing time, or walk-away time. In addition, the Liat per run workflow timing is identical to the influenza A/B and RSV workflow because of the three-analyte test.

**Table 2.**
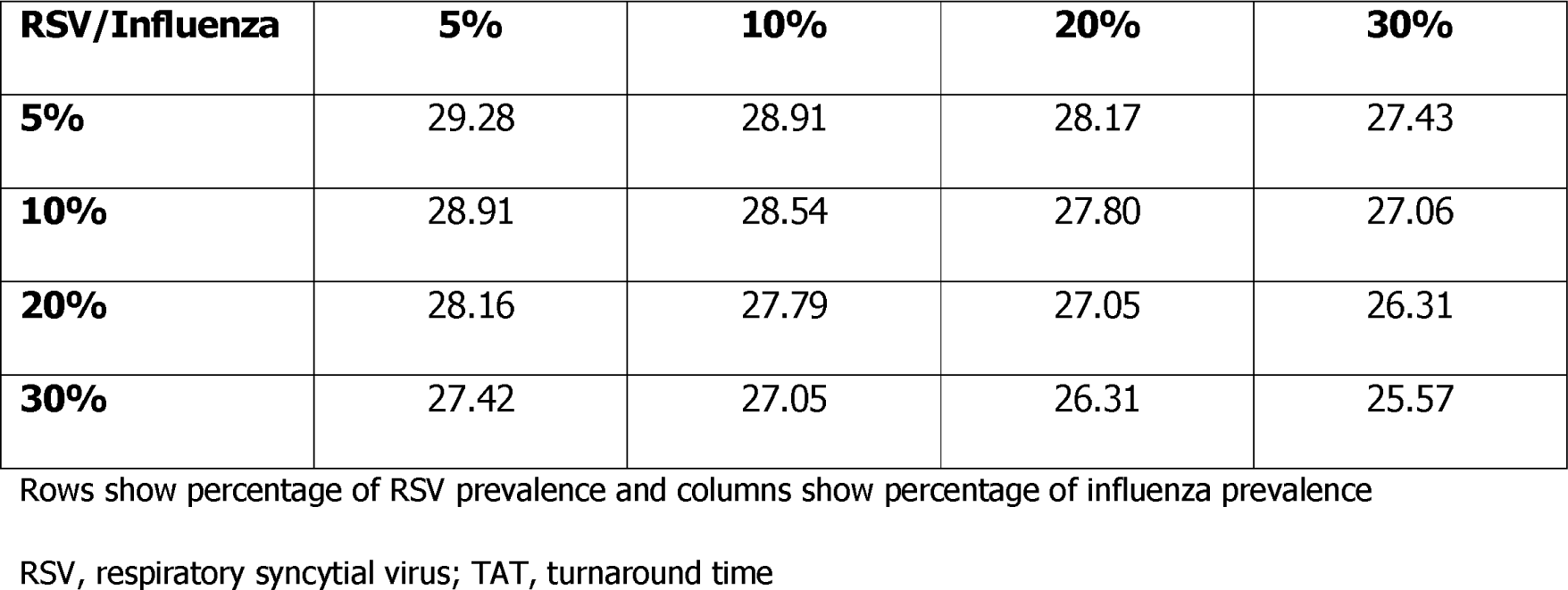
Expected TAT (minutes) for the ID Now influenza A/B and RSV assays (sequential workflow) by influenza and RSV prevalence

| **RSV/Influenza** | **5%** | **10%** | **20%** | **30%** |
| --- | --- | --- | --- | --- |
| **5%** | 29.28 | 28.91 | 28.17 | 27.43 |
| **10%** | 28.91 | 28.54 | 27.80 | 27.06 |
| **20%** | 28.16 | 27.79 | 27.05 | 26.31 |
| **30%** | 27.42 | 27.05 | 26.31 | 25.57 |

Rows show percentage of RSV prevalence and columns show percentage of influenza prevalence
RSV, respiratory syncytial virus; TAT, turnaround time

**Table 3.**
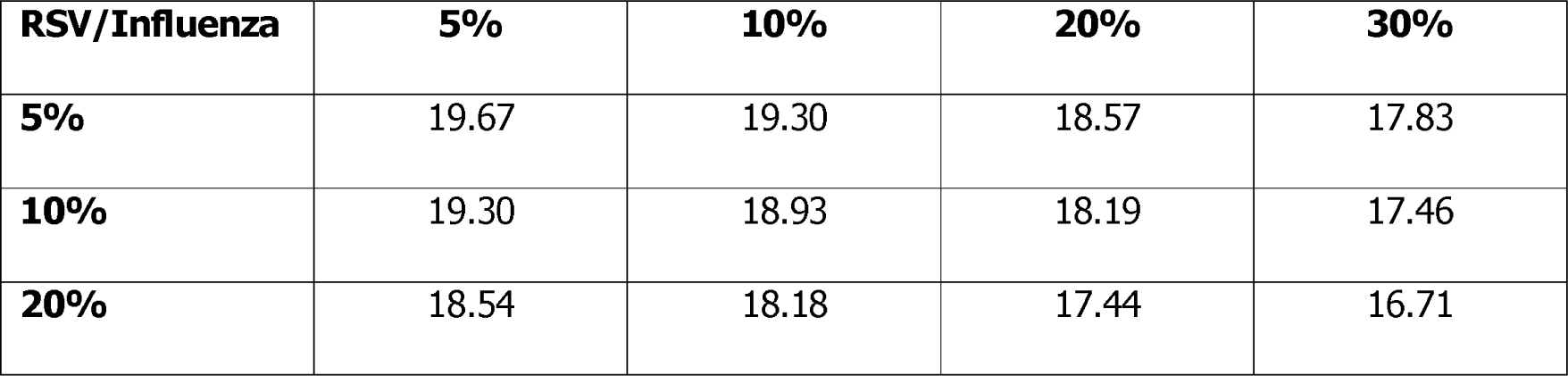
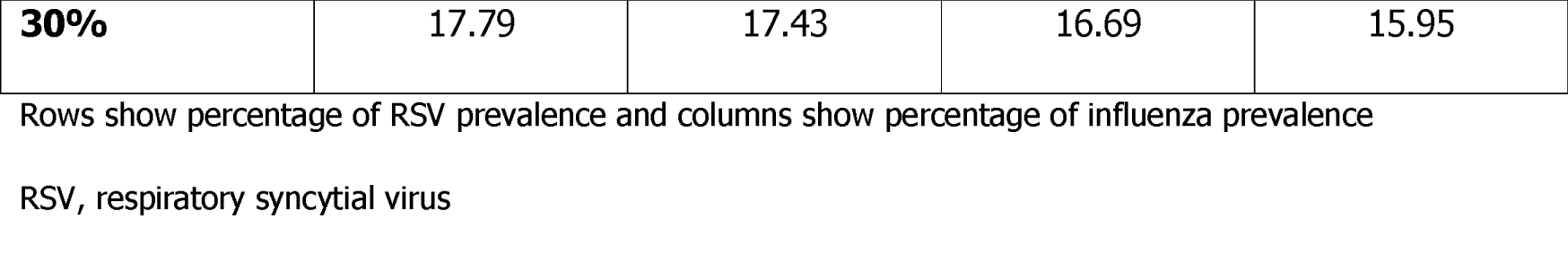
Expected walk-away time (minutes) for the ID Now influenza A/B and RSV assays (sequential workflow) by influenza and RSV prevalence

**Table 4.** Expected TAT (minutes) for the ID Now influenza A/B assay (sequential workflow) by influenza prevalence

| **Influenza prevalence** | **5%** | **10%** | **20%** | **30%** |
| --- | --- | --- | --- | --- |
| **Expected TAT (minutes)** | 14.74 | 14.37 | 13.63 | 12.88 |

TAT, turnaround time

### Discrepant results

There were discrepant results between the sequential and parallel workflow ID Now results for four samples in this study. Two samples (Sample IDs: 404-125 and 404-136) that were positive for influenza A under the sequential workflow were negative for influenza A/B and RSV under the parallel workflow, and two samples (Sample IDs: 404-128 and 404-133) that were negative for influenza A/B and RSV under the sequential workflow were positive for RSV under the parallel workflow. For the Liat system, test results for all 40 samples matched their original results for all tests during this study.

## DISCUSSION

The ID Now and Liat systems offer different workflow options for rapid diagnosis of influenza A/B and RSV in POC settings such as emergency rooms, urgent care centers, physician offices, and pharmacies. The median times reported in our study indicate that the ID Now sequential workflow has the longest TAT for influenza and RSV testing (up to 30 minutes) followed by the Liat system (21.6 minutes), and that the ID Now parallel workflow has the shortest TAT (15.1 minutes). The Liat has reduced staffing time (1.4 minutes compared with 5.9 (parallel)–9.3 (sequential) minutes) and increased walk-away time due to fewer manual steps compared with both ID Now workflows. The early call-out feature of the ID Now system enables shorter TATs for positive results compared with negative results (7.5 minutes vs 15.1 minutes).

When selecting the most appropriate diagnostic tool for influenza and RSV testing, it is critical to consider the potential benefits and drawbacks of each testing method. The ID Now and Liat systems each have their own potential benefits but these need to be considered in the context of the implementation scenario/setting. Specific factors to consider include location of instruments (e.g. whether they will be in a separate physician office lab, how much space is available for instruments), staffing and who will execute the tests (e.g. whether there will be a dedicated operator available who will stay with the instrument for the run, whether staff need to be multi-tasking and return to check the instrument). The Liat system requires less staffing time (sum of hands-on time and oversight time) (Figure 2) and fewer manual steps (Figure 1) than the ID Now system, providing advantages in settings without dedicated operators. The Liat system also has a consistent TAT, which is unaffected by pathogen prevalence. The consistent TAT may enable more effective planning because the time to results is fixed so that multi-tasking healthcare workers know when to return to the instrument to record the result. No manual intervention or operator wait times are required between test initiation and completion so there is not a risk of the instrument timing out. The Liat system is also multiplexed to detect influenza A/B and RSV during a single run so only one instrument is required to simultaneously test a sample for both respiratory viruses.

The ID Now system offers faster TAT compared with the Liat system when using a parallel workflow. A faster TAT could improve clinical workflow in settings like primary care with shorter appointment windows during busy winter respiratory seasons. To leverage the faster TAT of the ID Now, two instruments will be required per patient requiring results, so space to house instruments should be considered. Utilization of the ID Now system requires a high level of staffing time via direct interaction and vigilance (Figure 2) during each run so the availability and added cost of staff to operate the instruments should also be considered. The system will time out between steps 3 and 4 if the operation is not completed within 10 minutes.

The ID Now’s early call-out feature allows a very fast TAT for positive results. In our study, results were available at 6.4 minutes (minimum) to 9.6 minutes (maximum) for positive results and up to 15.8 minutes (maximum) for negative results. For a clinical site to leverage the workflow benefits of this 6–10-minute TAT, an operator would have to stay close to the instrument for up to 10 minutes or repeatedly return to check the instrument. As a result, this feature will provide the most utility in clinical settings with dedicated staff who stay with the instruments during the entire run or settings where the instruments are placed in the exam room with healthcare providers, who can monitor the test status during the exam. In a clinical setting, the majority of samples over the course of a respiratory infections season are negative; in the US 2018–2019 influenza season the peak influenza prevalence was ∼26%.[24] As a result, the time to receive negative results with the ID Now system (median 15.1 minutes for influenza) is a closer reflection of median TAT than the earlier time to positive results (median 7.7 minutes for influenza).

The discrepant results observed in our study with the ID Now system indicate that such inconsistencies are possible in a clinical setting and should be taken into consideration. This study provides robust, quantitative workflow data to aid the selection of a POC molecular test for influenza and RSV; however, there are several limitations that should be taken into account. This study was not conducted in a real-world clinical setting. The operators were trained laboratory technicians experienced with using the instruments; the results may have differed if the users were less experienced. Workflow was assessed using samples in batches of 10; in the real-world setting, a smaller or larger number may need to be tested during a working day. Furthermore, the samples used for testing were all positive for either influenza or RSV and had been previously frozen and thawed, so did not represent a typical set of clinical samples.

## CONCLUSIONS

The ID Now and cobas Liat systems offer different workflow options for emergency rooms, urgent care centers, physician offices, and pharmacies. When deciding which molecular test to implement, operational and workflow features should be considered as well as diagnostic performance/accuracy. Key considerations include the value of having both influenza and RSV results, staffing capacity, and instrument placement.

## Data Availability

Data are available upon reasonable request. The data are diagnostic workflow data derived from de-identified frozen specimens collected for routine clinical care in an excel spreadsheet. The data are available from Joanna Sickler and available until October 1, 2022. The data can be re-used subject to permission from Joanna Sickler. The study protocol is also available upon request.

## Key messages

- The use of POC NAAT tests for influenza and RSV in outpatient settings is increasing, offering the ability to improve patient management, infection control, and support antimicrobial stewardship
- This study provides data on workflow parameters (TAT, staffing time, and walk-away time) for the ID Now and cobas Liat systems, which can help to support the selection of the most appropriate influenza and RSV tests for different implementation scenarios
- The cobas Liat System offers a consistent TAT (21.3–22.0 minutes), with 1.4 minutes of staff time, and multiplexing capabilities so one test can provide influenza A, influenza B and RSV results
- The ID Now system offers TAT ranging 4.4–16.8 minutes, depending on analytes tested, workflow, and utilization of the early call-out feature, with staff requirements to stay with the instrument for 4.1–6.2 minutes per run

## Conflicts of interest

Stephen Young has participated in advisory boards for Roche Molecular Systems, Inc., Quidel, Inc., and Mesa Biotech, Inc. Jamie Phillips is an employee of Roche Diagnostics Corporation, Indianapolis, Indiana, USA. Shaowu Tang and Joanna Sickler are employees of Roche Molecular Systems, Inc., Pleasanton, California, USA. Sheena Chaudhuri was an employee of Roche Molecular Systems, Inc. at the time of the study.

## Acknowledgements

This study was conducted at TriCore Reference Laboratories (Albuquerque, New Mexico, USA) and funded by Roche Molecular Systems, Inc. (Pleasanton, California, USA). The workflow data was collected by Nexus. Medical writing support was provided by Papia Das. The authors thank Elissa Robbins for editorial review of the manuscript. COBAS and LIAT are trademarks of Roche. All other trademarks are the property of their respective owners.

## Notes

### Clinical Trial

Not applicable

### Funding Statement

This study was funded by Roche Molecular Systems, Inc. (Pleasanton, California, USA).

### Author Declarations

All relevant ethical guidelines have been followed and any necessary IRB and/or ethics committee approvals have been obtained.

Any clinical trials involved have been registered with an ICMJE-approved registry such as ClinicalTrials.gov and the trial ID is included in the manuscript.

